# Attempted Suicide and Criminal Justice System in a Sample of Forensic Psychiatric Patients in Malaysia

**DOI:** 10.1101/2021.06.30.21259822

**Authors:** Johari Khamis, Ravivarma Rao Panirselvam, Norhameza Ahmad Badruddin, Farynna Hana Ab Rahman, Lai Fong Chan

## Abstract

Criminalization of suicide attempts is an archaic barrier to suicide prevention. Globally, clinical profiles of prosecuted suicide attempters are an under-researched area. This retrospective study aims to describe the clinical profiles of individuals who were charged for attempted suicide and subsequently sent for criminal responsibility and fitness to plead evaluation in a forensic psychiatric unit in Malaysia from January 1, 2008, to December 31, 2019. We identified 22 cases who were mostly adult males (90.9%). Seventy-three percent have a psychiatric disorder. Mood disorders were more prevalent (32%) followed by psychotic disorders and substance use disorders. For most of these individuals, this was the first contact with any form of mental health services and 41% defaulted their treatment before arrest. This sample illustrates a vulnerable group who has been disengaged with mental healthcare. Future research is warranted to further investigate mechanisms that are effective in addressing unmet needs of persons in suicidal crisis as opposed to utilizing the criminal justice pathway.

## Introduction

The suicide rate in Malaysia is estimated to range from 6 – 8 per 100 000 (Armitage et al., 2015). Attempted suicide is a punishable offense in Malaysia, one of 25 countries documented to have specific statutes criminalizing suicide attempts (Mishara & Weisstub, 2016). Under Section 309 of the Penal Code (PC) (“Act 574,” 2018) an individual may be fined and/or jailed for up to one year if convicted. Current evidence suggest that such laws are not only ineffective in deterring suicide, but are likely counterproductive to help-seeking in suicide prevention (Mishara & Weisstub, 2016; Ng & Panirselvam, 2019). Preliminary findings from a systematic review showed that criminalisation of attempted suicide may increase, rather than decrease suicide rates. This effect seems more significant in countries with a lower Human Development Index (HDI) and among women in non-Muslim countries (Wu et al., 2020).

In relation to law, criminal responsibility, and fitness to plead are important considerations when the mental state of an alleged offender is of concern. Suicidal crisis is often associated with mental illness or states of significant mental distress (WHO, 2014). Therefore, the need for assessment of criminal responsibility and fitness to plead is particularly relevant in the event a person is criminally charged for attempting suicide. In Malaysian law, Section 84 of the PC applies to a person who is charged for an offence is not guilty by reason of insanity which is not knowing the nature or wrongfulness of action. The process of determining criminal responsibility and fitness to plead is provisioned by Section 342 of the Criminal Procedure Code (CPC) (“Criminal Procedure Code,” 2006) wherein an individual charged for an offense and is suspected to be of unsound mind during the alleged offence will be assessed in an approved government psychiatric hospitals with forensic psychiatry services under the provision of Section 22 of the Mental Health Act 2001. There are four such hospitals in Malaysia: Hospital Permai Johor Bahru, Hospital Bahagia Ulu Kinta, Hospital Sentosa Kuching, and Hospital Mesra Bukit Padang. Therefore, individuals who are charged under Section 309 of the PC, an evaluation can be ordered under Section 342 of the CPC to assess fitness to plead in the trial and assume criminal responsibility. The assessment findings will be conveyed as a report to the court which will be used as part of evidence in the trial. The outcome of such trials may be sentencing (fine and/or imprisonment), acquittal or detention in an approved government forensic hospital (if unable to assume criminal responsibility and/ or fitness to plead). The process is summarized in the Diagram 1.

**Diagram 1:**
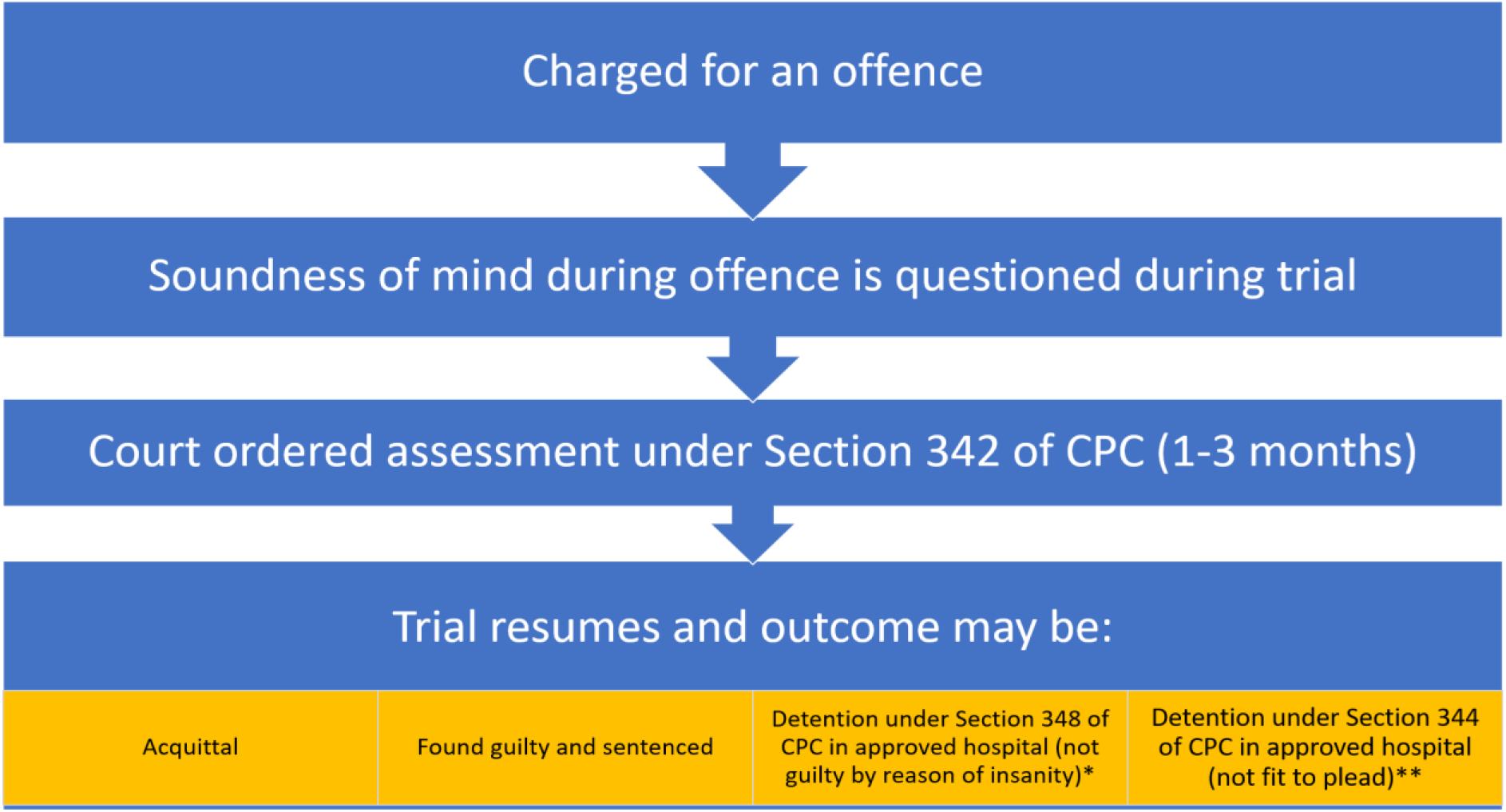
Assessment for Criminal Responsibility.

Globally, there is a lack of published evidence with regards to the implementation of criminal punishment towards suicide attempters in the context of criminal responsibility and fitness to plead. Anecdotal reports suggest that law enforcers have continued to apprehend and prosecute people who attempted suicide in the past decade. According to media reports, 11% of suicide attempters have been prosecuted from 2014-2018 in Malaysia (Jha, 2019; Shah, 2018). A qualitative study in Ghana showed that police officers were more inclined to facilitate help rather than arrest suicide attempters (Osafo et al., 2017). Prior to 2019 when suicide attempt was decriminalized in Singapore, 0.6% of reported attempted suicide were prosecuted on average yearly from 2013-2015 (*Penal Code Review Committee Report*, 2018). These findings seem to suggest that enforcement of such criminal charges is the exception rather than the current norm.

Nevertheless, not all persons charged for suicidal attempts are channeled for assessment of criminal responsibility and fitness to plead as this decision is based on the discretion of the magistrate presiding the hearing. In addition, it remains unclear whether the mental health needs of a person who has attempted suicide and subjected to legal proceedings are addressed. While calls for decriminalization remain a major priority of suicide prevention advocacy globally, literature examining the enforcement of the criminalization laws, especially those affected by it, are limited. Literature from Ghana have previously examined the sociodemographic, nature of attempts, and outcomes of people who criminally charged for suicidal attempts using media reports (Adinkrah, 2013) and qualitatively obtained the perspectives of stakeholders regarding their perspectives of criminalization of suicides (Hjelmeland et al., 2014; Osafo et al., 2017). Besides a recent review based on news and law casefiles (Thum Chern Chong & Chia, 2020), there is a paucity of published literature on this area in Malaysia. Furthermore, the characteristics of this population has yet to be clinically reviewed and is potentially a knowledge gap which if addressed, would build on our understanding with regards to the impact of prosecution and rationale for decriminalization within the context of low-middle income countries.

This retrospective study aims to describe the socio-demographic and clinical profiles of individuals who were charged under Section 309 of the PC for attempted suicide and subsequently sent for fitness to plead and criminal responsibility evaluation under Section 342 in Hospital Permai Johor Bahru (HPJB) from January 1, 2008, to December 31, 2019. The ethical approval for this study was obtained from the Medical & Research Ethics Committee (MREC), Ministry of Health Malaysia with the ID of NMRR-19-3348-51623. This study was investigator initiated and did not receive external funding.

## Methodology

This study employed a retrospective chart review design. Universal sampling of case records from the Records Unit of HPJB was performed for cases that were hospitalized within the study period of 1 January 2008 to 31 December 2019. Each case file was examined by the research team for the information required such as sociodemographic profile and clinical variables. Forensic data that was screened for study participant eligibility included data on court orders for assessment of criminal responsibility to Hospital Permai Johor Bahru under Section 342 of the CPC and charged under Section 309 of the PC from 1 January 2008 to 31 December 2019. The inclusion criteria were: adults 18 years and above, one of the charges of the offence must include Section 309 of the PC, detained under Section 342 of the CPC, and the case must be detained within the study time frame. Those who did not fulfil these criteria were excluded. All admissions within the study time frame that fulfilled the study criteria were included and analyzed. The data analysis was done using the SPSS version 22.

## Results

A total of 23 cases were admitted (to the forensic ward of HPJB) for an offence allegedly committed under Section 309 Penal Code, However, one case was excluded since the case was not detained under Section 342 of the CPC. Almost all the cases were male (90.9%). Two-thirds (72.7%) were Malaysian and almost half were Malay (45.5%) followed by Chinese (18.2%). Only one (4.5%) were of Indian and Indigenous descent each. Majority were from 30-39 years age group (mean age 37.7), received lower secondary school education, employed, and not married (Table 1)

**Table 1.** *Sociodemographic Profile* (N=22)

| Variables |  | n (%) |
| --- | --- | --- |
| Gender | Male | 20 (90.9) |
|  | Female | 2 (9.1) |
| Nationality | Malaysian | 16 (72.7) |
|  | Non-Malaysian | 6 (27.3) |
| Nationality/ ethnicity |  |  |
| Citizen | Malay | 10 (45.5) |
|  | Chinese | 4 (18.2) |
|  | Indian | 1 (4.5) |
|  | Indigenous Peoples | 1 (4.5) |
| Non-citizen<br>(Specified with nationality) | Bangladesh | 1 (4.5) |
|  | China | 1 (4.5) |
|  | Indonesia | 2 (9.1) |
|  | Nepal | 1 (4.5) |
|  | Sri Lanka | 1 (4.5) |
| Age (Mean=37.45) (SD=10.52) | 20-29 | 5 (22.7) |
|  | 30-39 | 9 (40.9) |
|  | 40-49 | 5 (22.7) |
|  | 50-59 | 2 (9.1) |
|  | 60-69 | 1 (4.5) |
| Education Level | Tertiary | 2 (9.1) |
|  | Upper Secondary | 3 (13.6) |
|  | Lower Secondary | 9 (40.9) |
|  | Primary | 5 (22.7) |
|  | No Formal Education | 2 (9.1) |
|  | Unknown | 1 (4.5) |
| Employment | Employed | 13 (59.1) |
|  | Unemployed | 9 (40.9) |
| Marital Status | Married | 7 (31.8) |
|  | Single | 9 (40.9) |
|  | Divorced/Separated | 6 (27.3) |

All court orders came from Magistrate Court, the designated court for cases charged with the 309 of the Penal Code. The majority of cases came from other states (63.6%), while Johor where Hospital Permai is located only account for 36.4%. These states are under the purview of Hospital Permai with regards to the court order for CPC. 16 (72.7%) cases were ordered for court assessment solely because of the 309 of the offences, the rest 6 (27.3%) have additional charges i.e., criminal intimidation, causing death by negligence, causing hurt, and not possessing valid travel documentation for foreign workers. Two-thirds did not have previous criminal offence.

All the cases were deemed fit to plead their charges and no person was readmitted to HPJB under Section 348 of the CPC (not guilty by reason of insanity and ordered for long term stay in an approved psychiatric hospital).

Unsoundness of mind is a Malaysian legal concept which origin can be traced back to the McNaughton Rule but based on the Indian Penal Code (Yeo et al., 2018). The defense of unsoundness of mind which is stated in Section 84 of the PC (“Act 574,” 2018) essentially holds the fundamental principle of criminal responsibility of a person upon committing a criminal offence. A successful plea on the defense of unsoundness of mind means at the time of the alleged offence, the accused must have suffering of unsound mind which in turn impaired the capacity to know the nature of the offence and its either wrong or contrary to the law. The term ‘unsoundness of mind’ was not defined by the PC.

Fitness to plead is also a legal concept used in the Malaysian Criminal Law. Unlike unsoundness of mind, it was not codified in any statute but almost always required to be made in Section 342 of the CPC of court orders for psychiatric assessment. For the purpose of determining the fitness to plead of the accused, the psychiatrist applies the Pritchard Criteria (Leong et al., 2021) which include the ability to understand the nature of the charge and the possible consequences of a finding of guilt, to instruct his legal counsel, and to understand the evidence against him.

Lethality is characterized as the need for hospital admission. One-third of the cases attempted suicide by using sharp objects (40.9%), followed by hanging and falling from height, both 22.7%. More than half attempted suicide at private places. Notably, three of the cases (13.6%) occurred while incarcerated in prison for other offences. Suicidal intent was reported in more than half of the population, while the remaining did not report any intent i.e., charged for attempts of self-harm without suicidal intent or the intent was not elicited by the police. Forty-point nine percent of the cases reported negative life events surrounding the event of the attempt. Another 13.6% had used substances at the time.

The most common psychiatric disorder was mood disorders (31.8%), followed by substance use disorders including alcohol use disorder (18.2%). However, those with previously diagnosed mental illness, 90% (n=9) of the population had defaulted treatment. More detailed results were shown in Table 5. Twelve cases (54.5%) did not have a history of psychiatric disorder as they had no previous contact with mental healthcare. Of these, seven (58.3%) were found to have a diagnosable mental health disorder during their assessment.

**Table 2.** *Forensic Variable*s (N=22)

| Variables |  | n (%) |
| --- | --- | --- |
| Court | Magistrate | 22 (100) |
| State | Johor | 8 (36.4) |
|  | Kelantan | 2 (9.1) |
|  | Melaka | 3 (13.6) |
|  | Negeri Sembilan | 1 (4.5) |
|  | Pahang | 7 (31.8) |
|  | Terengganu | 1 (4.5) |
| Additional Charge | No Additional Charges | 16 (72.7) |
|  | Criminal intimidation | 1 (4.5) |
|  | Causing death by negligence | 1 (4.5) |
|  | Grievous hurt | 2 (9.0) |
|  | Immigration offences | 2 (9.1) |
| Past Forensic History | Yes | 8 (36.4) |
|  | No | 14 (63.6) |

**Table 3.**
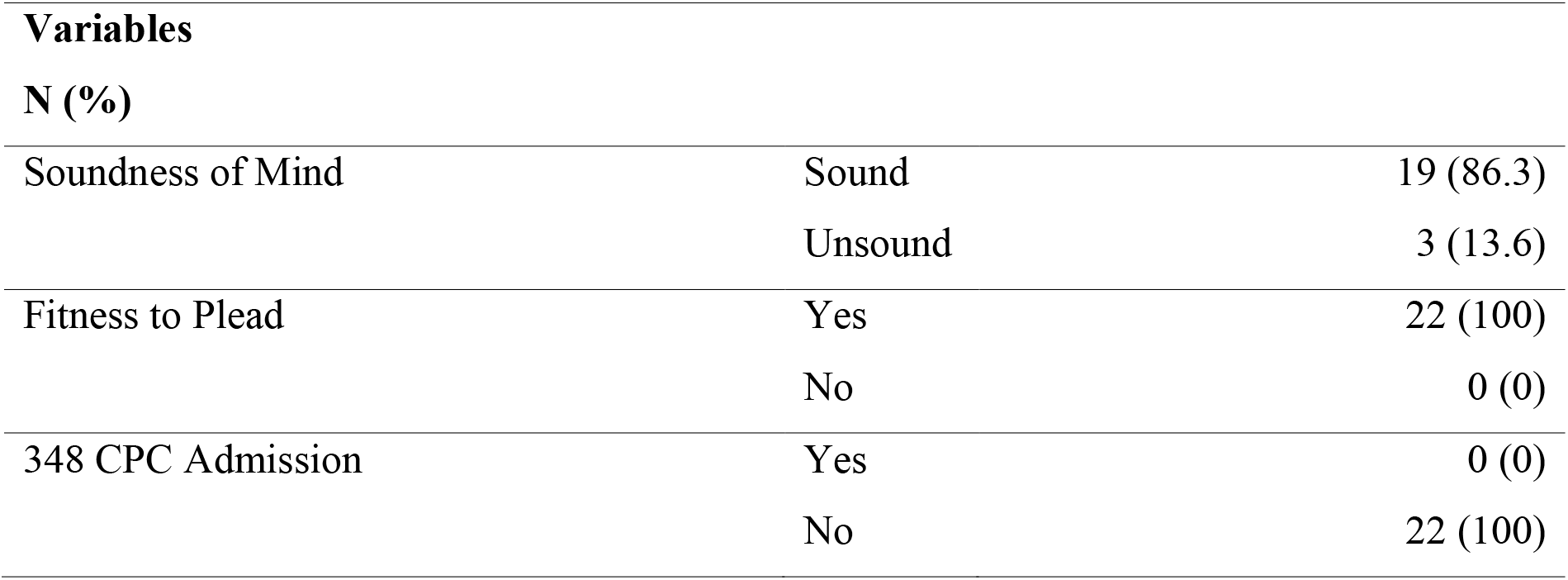
Criminal Responsibility.

**Table 4.**
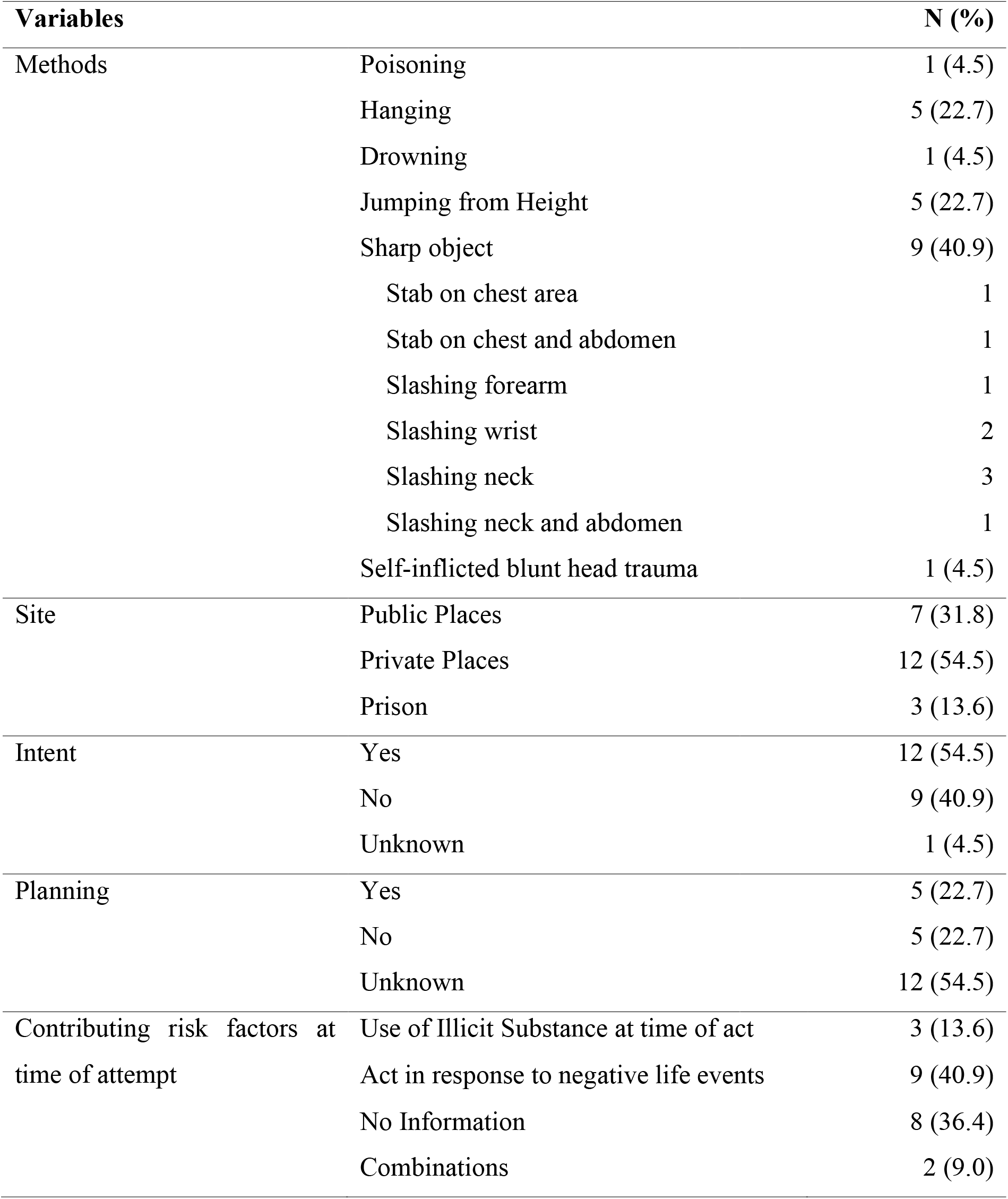
Suicide Attempt Variable.

**Table 5.** Clinical Variables.

| **Variables** |  | **n (%)** |
| --- | --- | --- |
| Diagnosis | No Mental Disorder | 6 (27.3) |
|  | Psychotic Disorders | 5(22.7) |
|  | Mood Disorders | 7(31.7) |
|  | Substance Use Disorder (including alcohol) | 4(18.2) |
| Previous Psychiatric History | Yes | 9 (40.9) |
|  | No | 12 (54.5) |
|  | Unknown | 1 (4.5) |
| Disengagement from Service<br>(defaulted from previous<br>treatment) | Yes | 9 (40.9) |
|  | No | 1 (4.5) |
|  | Not applicable | 12 (54.5) |

## Discussion

From 2008 until 2019, Hospital Permai, Johor Bahru recorded 2,810 admissions to its forensic psychiatry unit. From that figure, 23 admissions were related to suicide attempts, excluding one admission, the other 22 cases were admitted under Section 342 of the CPC after they were charged for suicide attempt under Section 309 of the PC. These were only accounted for 0.78% of all the admissions.

Despite the small proportion of cases relative to the total caseload, the results indicate this law is indeed enforced bearing in mind not all who were charged for attempting suicide in the catchment area of HPJB were channeled for assessment under Section 342 of the CPC. There certainly exists an iceberg phenomenon with regards to the actual number of people who were charged under Section 309 of the PC as the total records were not available in the public domain. Nevertheless, the information obtained in this study provides an accurate and objective clinical assessment of the persons charged for suicidal attempts.

The authors summarize the usual profile of a person is a single male in his thirties likely to be Malaysian Malay who has lethally attempted suicide. This data is not surprising as suicide rates were found to be similar in Muslim vs. non-Muslim populations (Lawrence et al., 2016; Lester, 2006). The authors considered lethality as medically serious suicidal behavior (WHO, 2014). This person who attempts suicide is likely to be employed, less than nine years of schooling and has an untreated mental illness especially a mood disorder, and previously disengaged from services. Similar low socioeconomic profiling was noted in a Ghanaian study (Adinkrah, 2013).

Notably, these attempts occur in private with intent and surrounding negative life events. Three cases happened in correctional institutions (prison facilities). In regard to prisoners, it has been found nearly 20% of prisoners engaged in suicidal attempts (Larney et al., 2012) and found near-lethal attempts were associated with psychiatric disorders (Rivlin et al., 2010). In short, it can be summarized that this population is vulnerable (Dahlgren & Whitehead, 1991; Waisel, 2013).

This ties in with how our population came into contact with healthcare services i.e., the criminal justice pathway. While we cannot form strong associations within the population that is charged under Section 309 of the PC and a much larger population of people who self-harm due to the paucity of published data, this data makes us question the nature of care especially support after a suicidal crisis. Section 342 of the CPC provisions for assessment and treatment is ancillary when in fact the Mental Health Act 2001 (MHA 2001) (“Mental Health Act 2001,” 2001) which provisions for treatment for suicidal patients either voluntary or involuntary is not utilized by the police. We wonder if the longer duration of assessment for criminal responsibility and fitness to plead in a secure set-up ranging from one to three months is helpful for a person’s mental health. In our data, three cases were found to be of unsound mind. In legal terms, this could mean that they were found to be not guilty because of insanity (NGRI) according to Section 84 of the PC if the psychiatrist opinion were accepted by the court. The authors wonder if they were acquitted, fined or incarcerated as they were not returned to the said forensic facility under Section 348 of CPC and if they are receiving the appropriate mental healthcare.

Reflecting on the process that the individuals undergo as part of criminal responsibility assessment, we can postulate those individuals such as those in the study sample spend a significant duration of time in the criminal justice pathway both before and after assessment. This prolonged timeline may worsen the risk of suicidal attempt especially if the person is not provided access to necessary supports within the system. While treatment and mental healthcare is provided in tandem with the assessment, the continuity of care remains of the question despite clinical recommendations are made as part of the court report and discharge referral for such individuals. This when superimposed on the vulnerability of this population especially with regards to disengagement and access to care, the risk of poor mental health and future suicide attempts became a serious concern (WHO, 2014).

Whether being charged provides any preventive efforts of future suicide attempts in the persons charged in this population remains unknown as there are no surveillance systems in place post-discharge. The impact this law would have even in our small sample is worrying where individuals 90% of those who were previously diagnosed to have mental disorders had disengaged from services and how this would affect their future health-seeking patterns. It must be also noted that more than half who did not have a diagnosable psychiatric disorder before admission were diagnosed with one during the assessment. A previous attempt is a significant risk factor for a future attempt (WHO, 2014). Decriminalization has not been found to increase rates of suicide in a review (IASP, 2020). Researchers opined that suicide rates would decline and initially increased rates of reporting as fear of persecution and concealment is lifted. The rates of “undetermined death” fell significantly following decriminalization (Osman et al., 2017). The International Association of Suicide Prevention (IASP) holds the standing that criminalization undermines suicide prevention and impedes vulnerable individuals from accessing resources in preventing suicides. Furthermore, IASP recommends that decriminalization will reduce stigma and barriers to care, improve engagement in suicide prevention activities and care, along better surveillance of suicidal behavior.

In our context and the vulnerability of our population, we raise further apprehension of whether utilizing the criminal justice pathway is cost-effective in providing care with limited benefit. In a broader aspect, Section 309 of the PC and the criminal justice pathway does not delineate non-suicidal self-injury from a suicidal attempt, which further raises concern for this pathway when a defined and holistic pathway of care through the Mental Health Act 2001 (“Mental Health Act 2001,” 2001) exists. Finally, we concur with IASP that people who attempt suicide have a reduced capacity to make an informed decision due to their mental illness, substance abuse, or extreme crisis. Hence, suicidal behavior is unlikely deterred by the criminalization of suicide attempts and in fact, it can worsen suicidal risk in people who are incarcerated without the necessary support.

Our study’s strength is being the quality of the data wherein clinical profiling of the person being charged was documented by a trained multidisciplinary forensic psychiatry team as opposed to accuracy and biases of second or third person accounts in media or other stakeholders. We would highlight systemic limitations especially follow-up of the sample population remains unknown especially if they were sentenced, received follow-up care and further clinical outcomes.

## Conclusion

Suicides happen in complex situations and to people in heightened distress. Consistently, evidence has shown no benefit of criminalizing suicides which is a public health crisis. Our data, if at all, has shown vulnerable individuals who otherwise should have received support and mental healthcare first are charged under Section 309 of the PC. We recommend future work in this field to evaluate the economic costs of this process comparing that through the MHA 2001 pathway (“Mental Health Act 2001,” 2001), seek to describe the lived experience of individuals who are prosecuted on this charge and support efforts for the decriminalization of suicidal attempts.

## Data Availability

Public access data not available.

## References

Act 574, 52; 178 § 84; 309 (2018). https://agc.gov.my/

Adinkrah, M. (2013). Criminal prosecution of suicide attempt survivors in Ghana. International journal of offender therapy and comparative criminology, 57(12), 1477–1497. https://doi.org/ https://doi.org/10.1177/0306624X12456986

Armitage, C. J., Panagioti, M., Rahim, W. A., Rowe, R., & O’Connor, R. C. (2015). Completed suicides and self-harm in Malaysia: a systematic review. General hospital psychiatry, 37(2), 153–165. https://doi.org/10.1016/j.genhosppsych.2014.12.002

Criminal Procedure Code, § 342 (1) (2006). https://agc.gov.my/

Dahlgren, G., & Whitehead, M. (1991). Policies and strategies to promote social equity in health. Background document to WHO-Strategy paper for Europe.

Hjelmeland, H., Osafo, J., Akotia, C. S., & Knizek, B. L. (2014). The law criminalizing attempted suicide in Ghana. Crisis. https://doi.org/10.1027/0227-5910/a000235

IASP. (2020). The Decriminalisation Of Suicidal Behaviour. International Association for Suicide Prevention. Retrieved 12/05/2021 from https://www.iasp.info/decriminalisation/

Jha, P. (2019, 26 June 2019). Will Malaysia End Its Archaic Suicide Law? The Diplomat. https://thediplomat.com/2019/06/will-malaysia-end-its-archaic-suicide-law/

Larney, S., Topp, L., Indig, D., O’Driscoll, C., & Greenberg, D. (2012, 2012/01/06). A cross-sectional survey of prevalence and correlates of suicidal ideation and suicide attempts among prisoners in New South Wales, Australia. BMC Public Health, 12(1), 14. https://doi.org/10.1186/1471-2458-12-14

Lawrence, R. E., Oquendo, M. A., & Stanley, B. (2016). Religion and suicide risk: a systematic review. Archives of Suicide Research, 20(1), 1–21. https://doi.org/10.1080/13811118.2015.1004494

Leong, K. K., Chai, Y. C., Johari, K., Hosni, N. S., & Badruddin, N. A. (2021). On Fitness to Plead: When the Accused is Speech-less. Malaysian Journal Of Psychiatry, 30(1).

Lester, D. (2006). Suicide and Islam Archives of Suicide Research, 10(1), 77–97. https://doi.org/10.1080/13811110500318489

Mental Health Act 2001, (2001). https://agc.gov.my/

Mishara, B. L., & Weisstub, D. N. (2016). The legal status of suicide: A global review. International journal of law and psychiatry, 44, 54–74. https://doi.org/10.1016/j.ijlp.2015.08.032

Ng, Y. P., & Panirselvam, R. R. (2019). The Endgame of Section 309? An Appeal for Decriminalisation of Suicide. Malaysian Journal Of Psychiatry, 28(1), 1–6.

Osafo, J., Akotia, C. S., Quarshie, E. N.-B., Boakye, K. E., & Andoh-Arthur, J. (2017). Police views of suicidal persons and the law criminalizing attempted suicide in Ghana: A qualitative study with policy implications. Sage open, 7(3), 2158244017731803. https://doi.org/10.1177/2158244017731803

Osman, M., Parnell, A. C., & Haley, C. (2017). “Suicide shall cease to be a crime”: suicide and undetermined death trends 1970–2000 before and after the decriminalization of suicide in Ireland 1993. Irish Journal of Medical Science (1971-), 186(1), 201–205. https://doi.org/10.1007/s11845-016-1468-9

Penal Code Review Committee Report. (2018). https://www.reach.gov.sg/-/media/reach/old-reach/2018/public-consult/mha/annex--pcrc-report.ashx

Rivlin, A., Hawton, K., Marzano, L., & Fazel, S. (2010). Psychiatric disorders in male prisoners who made near-lethal suicide attempts: case–control study. The British Journal of Psychiatry, 197(4), 313–319. https://doi.org/10.1192/bjp.bp.110.077883

Shah, A. (2018, August 26, 2018). Over 2,000 Suicide Cases in the Past Four Years. New Straits Times. https://www.nst.com.my/news/exclusive/2018/08/404930/exclusive-over-2000-suicide-cases-past-four-years-nsttv

Thum Chern Chong, & Chia, M. P. F. (2020). Reconciliation of Law and Medicine with Reference Made to Section 309 Penal Code. Malayan Law Journal, 5(lxxxi).

Waisel, D. B. (2013). Vulnerable populations in healthcare. Current Opinion in Anesthesiology, 26(2), 186–192. https://doi.org/10.1097/ACO.0b013e32835e8c17

WHO. (2014). Preventing suicide: A global imperative. World Health Organization.

Wu, K. C.-C., Cai, Z., Chang, Q., Chang, S.-S., Yip, P. S. F., & Chen, Y.-Y. (2020). Criminalization of Suicide and Suicide Rates in the World. https://ssrn.com/abstract=3564402

Yeo, S., Morgan, N., & Chan, W. C. (2018). Criminal law in Malaysia and Singapore. LexisNexis. https://ink.library.smu.edu.sg/sol_research/3061

